# Association between Clauss fibrinogen concentration and ROTEM® FIBTEM A5 in women with postpartum haemorrhage: validation of cut-off points

**DOI:** 10.1101/2023.10.12.23296942

**Authors:** Ada Gillissen, Bahar Golyardi, Thomas van den Akker, Camila Caram-Deelder, Dacia D.C.A. Henriquez, Kitty W.M. Bloemenkamp, Johannes J. Duvekot, Moniek P. M. de Maat, Jeroen Eikenboom, Johanna G. van der Bom

## Abstract

**Objective:** Point-of-care tests like FIBTEM A5 have been proposed to guide the treatment of patients who might have low fibrinogen concentrations (≤2 g/L). The aim of this study was to describe fibrinogen concentrations according to previously proposed FIBTEM A5 cut-off points in blood samples collected from women during postpartum haemorrhage.

**Design and setting:** Prospective multicentre cohort study

**Population:** 511 women sustaining postpartum haemorrhage. A total of 637 blood samples were drawn during haemorrhage.

**Methods:** Clauss fibrinogen concentrations and ROTEM® FIBTEM A5 values were studied to assess the diagnostic properties of previously proposed FIBTEM A5 cut-off points for the detection of low fibrinogen concentrations.

**Main outcome measures:** Youden index, sensitivity and specificity

**Results:** Of 511 women with a median total volume of blood loss of 1500 mL (IQR 1200 to 2000) 31 women (6%) developed Clauss fibrinogen concentrations below 2 g/L. Using FIBTEM A 5 cut-off of ≤ 7 mm: 48% of cases with Clauss fibrinogen ≤2 g/L were missed (FIBTEM A5 > 7mm), and of the 28 samples with FIBTEM A5 ≤ 7mm, 12 (43%) samples had Clauss fibrinogen >2 g/L. Using FIBTEM A5 cut-off of ≤12mm: 13% of cases with Clauss fibrinogen ≤2 g/L were missed and of the 145 samples with a FIBTEM A5 ≤12 mm, 118 had Clauss fibrinogen >2 g/L, resulting in false positive selection of 81% of women. Using FIBTEM A5 ≤ 15 mm: 97% (30/31) of the samples with Clauss fibrinogen ≤2 g/L were accurately selected; yet 89% (248/278) of samples that were selected had a fibrinogen concentration of >2 g/L. Based on the Youden index, the optimal cut-off point in our cohort was a FIBTEM A5 of 12mm with sensitivity 87% and specificity 81%.

**Conclusions:** Our findings suggest that if FIBTEM A5 lower than 12 mm would have been used to detect women with fibrinogen concentrations below 2 g/L in order to treat them with fibrinogen concentrate, 87 % of the women with fibrinogen below 2 g/L would correctly have received fibrinogen. However, most women (81%) receiving fibrinogen concentrate would not have needed it, because they had plasma fibrinogen concentrations above 2 g/L.

**Funding:** no funding was obtained to conduct this research

## Introduction

Postpartum haemorrhage remains one of the leading maternal health problems worldwide^1, 2^. Although in most women the primary cause is obstetric, acquired haemostatic impairment may aggravate bleeding^3, 4^. From previous studies, a low fibrinogen concentration emerges as the earliest predictor of progression towards severe postpartum haemorrhage^5, 6,7–10^. By timely detection of low fibrinogen concentrations, targeted haemostatic therapy may be administered to restore adequate concentrations of fibrinogen. The Clauss fibrinogen assay is the standard coagulation test to assess fibrinogen concentration. Its downside is a turn-around time of up to 60 minutes, rendering this test unsuitable for clinical decision making in the acute setting^11^. Point-of-care devices like ROTEM^®^ thromboelastometry can detect changes in the coagulation system within ten minutes from blood sampling^4^. In trauma, cardiac and liver surgery, thromboelastometry is increasingly used to predict bleeding, diagnose fibrinogen deficiency and guide fibrinogen administration. Evidence is emerging that ROTEM-guided transfusion strategies may reduce the need for blood products and bleeding- associated morbidity^12^. ROTEM^®^ devices are becoming more widely available in maternity wards, where treatment is generally dependent on local algorithms. Several studies in women sustaining postpartum haemorrhage found that fibrinogen concentrations below 2 g/L were associated with progression towards more severe postpartum haemorrhage^5–9, 13^. The ROTEM^®^ FIBTEM A5 assay, available within 7 to 10 minutes after sampling, may provide a quantitative assessment of the plasma concentration of fibrinogen of women during postpartum haemorrhage and has been established as an early biomarker for progression towards more severe postpartum haemorrhage^4, 13, 14^. However, the diagnostic properties of FIBTEM A5 to accurately identify Clauss fibrinogen concentrations ≤2 g/L remain unclear. In a previous trial comparing administration of fibrinogen concentrate with placebo in women with postpartum haemorrhage, a FIBTEM A5 cut-off of 15 mm was used and no difference was observed between groups with regard to number of units of red blood cells, plasma, cryoprecipitate and platelets transfused^15^. An earlier study in women with postpartum haemorrhage suggested a FIBTEM A5 value of 6 mm as the cut-off point that correlates best with Clauss fibrinogen below 2 g/L. To create reliable ROTEM^®^- based treatment algorithms for use during postpartum haemorrhage, the properties of the FIBTEM A5 test to detect Clauss fibrinogen concentrations below 2 g/L among women at risk of severe outcome of postpartum haemorrhage needs to be established.

The aim of this study was to describe fibrinogen concentrations according to previously proposed FIBTEM A5 cut-off points in blood samples collected from women suffering persistent postpartum haemorrhage.

## Methods

### Design and study population

We studied women who had been included in the TeMpOH-2 (Towards better Prognostic and Diagnostic strategies for Major Obstetric Haemorrhage) study, a prospective cohort of pregnant women in the Netherlands between February 2015 and April 2018. Women were recruited during pregnancy at the outpatient clinics and maternity wards of three participating hospitals: the Leiden University Medical Center, the Erasmus Medical Centre Rotterdam and the Zwolle Isala Clinics.

Included women were monitored for the occurrence of postpartum haemorrhage (≥1000 mL blood loss within 24 hours from childbirth) and followed until discharge from hospital. All subjects were treated according to the Dutch regulations for prevention and management of postpartum haemorrhage^16^. Blood samples were drawn from women in case they developed postpartum haemorrhage. Up to three samples per woman were sampled, with the first sample drawn at a blood loss between 1000-1500mL, but in some cases pragmatically drawn from 800mL of blood loss at time of achieving intravenous access. Subsequent samples were drawn in case of an additional volume of blood loss of 1000-1500mL. Both a ROTEM® FIBTEM assay and fibrinogen concentration assessed by the Clauss method were performed. A Clauss fibrinogen concentration of ≤2g/L was considered to be a low fibrinogen level. FIBTEM cut-off points of 7, 12 and 15 mm that were used in previous studies as an equivalent of a low fibrinogen (≤2g/L) were compared to Clauss fibrinogen concentrations. ROTEM thromboelastometry results were not available to treating clinicians. During the inclusion period of the TeMpOH-2 study, the ROTEM® Delta was replaced by the ROTEM® Sigma device. On the ROTEM® Sigma device, measurements are performed in a fully automated manner. In one of the study sites measurements were performed simultaneously with both devices^17^. Approval for the study was obtained from the Ethical Committee of the Leiden University Medical Centre (P13.246) and from the institutional review board of each participating hospital. The study was registered at ClinicalTrials.gov (NCT02149472). The ethical committee provided the possibility to ask women for verbal informed consent during early postpartum haemorrhage in case they had not yet been included in the study during pregnancy. The present analysis was restricted to data from women who provided written informed consent for their data to be used in the study. Women below 18 years of age or with a gestational age below 24 weeks at the time of birth were excluded. Women with coagulation disorders or who used anticoagulants were not excluded.

### Data collection

Information on maternal and obstetric characteristics was collected by well-trained research nurses who performed comprehensive chart reviews. Data were recorded from medical files available at the maternity ward for the following parameters: maternal age at the time of birth, parity, gestational age, mode of birth, presence of pre-eclampsia or HELLP syndrome, presence of a coagulation disorder, anticoagulant use and total volume of blood loss, primary cause of major obstetric haemorrhage, abnormal placentation, shock, volume and timing of administration of fluids and blood products, performance and timing of surgical and haemostatic interventions, and consecutive measurements of blood loss until cessation of bleeding. Blood loss was measured by weighing gauzes and all other soaked materials and by use of a collector bag and suction system in the operating theatre.

### Handling of ROTEM® devices and measurements

In two of the participating hospitals, the ROTEM® devices were positioned at the laboratory and samples were handled by laboratory staff. In the other hospital, the devices were positioned in a utility room equipped to house laboratory devices at the maternity ward. Here, ROTEM® measurements were performed by well-trained study personnel including research nurses and well-trained clinical midwives. In case measurements could not be started immediately, citrated blood samples were stored and handled at 37°C. Results were only taken into consideration when measurements had started within 4 hours after blood sampling, since sample stability up to 4-6 hours has been confirmed in previous studies^18, 19^. Blood draw was performed by venepuncture using a 21 -gauge blood collection needle or following insertion of a peripheral vein cannula. Blood was collected in vacuum tubes (BD Vacutainer® Citrate tubes 3.2% and BD Vacutainer® spray-coated K2EDTA tubes), always discarding the first 3 mL of blood. Citrated tubes were collected before EDTA tubes. Visual inspection was conducted to verify if the minimum acquired volume of blood had been collected. If not, the tube was discarded. In case samples were not handled directly at the maternity ward, tubes were sent to the laboratory by pneumatic tube system transport. Blood samples for Clauss fibrinogen assays were handled immediately after arriving at the laboratory and centrifuged for 10 minutes at 2700 g at a temperature of 20°C. External quality control for Clauss fibrinogen, APTT and PT and (in two of three participating hospitals) ROTEM® parameters was secured by participation in the international External Quality Assessment Programme (EQAP) by ECAT. Internal quality control was performed weekly by using the quality control reagents ‘ROTROL N’ (normal control) and ‘ROTROL P’ (abnormal control) provided by the manufacturer. Also, daily, quarterly and yearly maintenance on the devices was carried out according to the manufacturer’s instructions. Single use reagents were used in the ROTEM® Delta device.

### Statistical analyses

Characteristics of women are reported using descriptive statistics. Clauss fibrinogen and ROTEM parameters are presented as median and interquartile ranges because of their non-Gaussian distribution. In case of multiple samples per woman these were all taken into account in the analysis. All ROTEM^®^ parameters were verified by visually inspecting corresponding temograms for normal layout and by checking all parameters for error codes. Incorrect values were excluded from analysis. Spearman’s rank correlation coefficients (r_s_) were assessed for the correlation between ROTEM® FIBTEM values of both devices and fibrinogen concentration as obtained by Clauss assay^20^. Sensitivity, specificity, positive and negative predictive value and the area under the receiver operator curve (AUC’s) were calculated to assess the diagnostic properties of FIBTEM A5 to discriminate women with and without a fibrinogen concentration of ≤2g/L. The cut-off with the highest sensitivity and specificity was determined by use of the Youden index, which defines the maximum potential effectiveness of a test^21^. A complete case analysis was performed, data were not imputed.

## Results

### Women characteristics

Over the three-year inclusion period, 17203 women gave birth in the participating hospitals. Of all women, 1605 suffered postpartum haemorrhage defined as a volume of blood loss of ≥1000 mL within 24 hours from childbirth and 591 women agreed to participate in the study. For 511 women, valid corresponding measurements of fibrinogen and FIBTEM A5 were available resulting in 637 samples (Figure 1). Baseline characteristics are reported in Table 1. Women were on average 32 years of age (interquartile range (IQR) 28 to 35), gave birth at a median gestational age of 39.4 weeks (IQR 38.0 to 40.6) and in 23% by caesarean section. Median total volume of blood loss was 1500mL (IQR 1200 to 2000) and the most prevalent causes of haemorrhage were atony and/or retained placenta in 76% of women. One sample was available for 397 women, a second sample for 102 and a third sample for 12 women.

**Figure 1:**
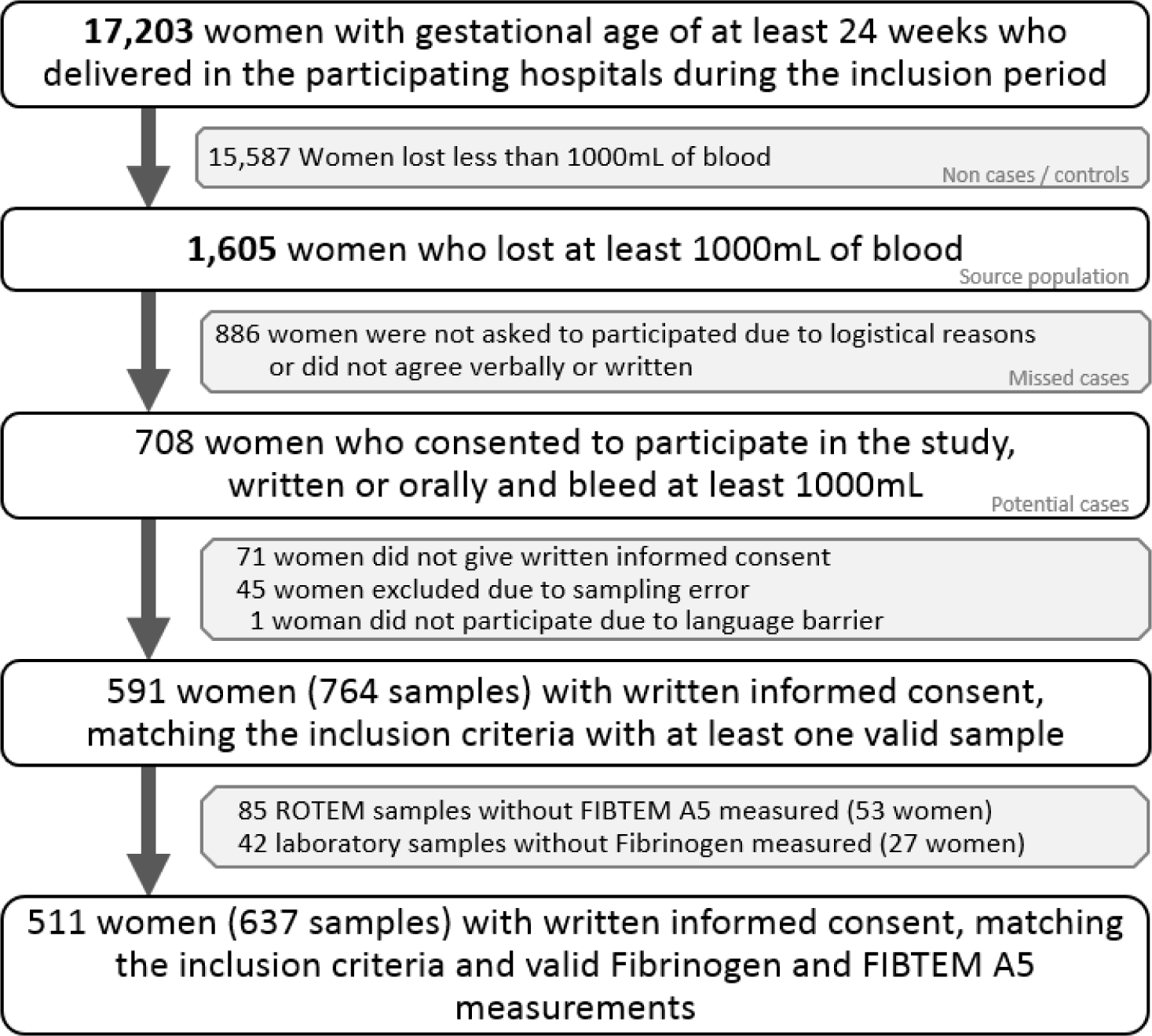
inclusion flowchart

**Table 1:** Women characteristics.

|  | Women |
| --- | --- |
| **Maternal characteristics** | n =511 |
| Age in years | 32 (28 to 35) * |
| BMI in kg/m<sup>2</sup> | 24 (22 to 28) * |
| Ethnicity, Caucasian | 86% |
| Gestational age in weeks | 39.4 (38.0 to 40.4) * |
| Nulliparity | 51% |
| **Risk factors PPH** |  |
| Pre-eclampsia/HELLP | 4% |
| Anti-coagulant use | 3% |
| Known coagulation disorder |  |
| Multiple pregnancy | 9% |
| **Mode of birth** |  |
| Vaginal | 77% |
| Caesarean section | 23% |
| **Primary cause of bleeding** |  |
| Uterine atony | 35% |
| Retained placenta or remnants of placental tissue | 41% |
| Surgical bleeding | 9% |
| Genital tract trauma | 9% |
| Placenta praevia | 2% |
| Pathological ingrowth of placenta | 1% |
| Placental abruption | 2% |
| Uterine rupture | 1% |
| Coagulopathy | 0% |
| **Bleeding** |  |
| Total volume of clear fluids (L) | 1.0 (0.5 to 2.0) * |
| Total units of blood products (n) | 0 (0 to 0) * |
| Total volume of blood loss (n) | 1500 (1200 to 2000) * |
| **Samples** |  |
| **Sample 1000-1500mL** | n=412 |
| Blood loss at sampling time (L) | 1.0 (0.9 to 1.2) * |
| Units of RBC received at sampling time (n) | 0.0 (0.0 to 0.0) * |
| Fluids received at sampling time (L) | 1.0 (0.0 to 2.0) * |
| **Sample 1500-2500mL** | n=185 |
| Blood loss at sampling time (L) | 1.8 (1.5 to 2.0) * |
| Units of RBC received at sampling time (n) | 0.0 (0.0 to 0.0) * |
| Fluids received at sampling time (L) | 2.0 (1.0 to 4.0) * |
| **Sample >2500mL** | n=40 |
| Blood loss at sampling time (L) | 2.7 (2.5 to 3.4) * |
| Units of RBC received at sampling time (n) | 0.0 (0.0 to 2.0) * |
| Fluids received at sampling time (L) | 3.0 (2.0 to 6.0) * |
| **Total** | n=637 |
| Blood loss at sampling time (L) | 1.2 (1.0 to 1.6) * |
| RBC received at sampling time | 0.0 (0.0 to 0.0) * |
| Fluids received at sampling time (L) | 1.0 (1.0 to 3.0) * |

\* All values in the table reported in this format are median and (IQR)

### Clauss Fibrinogen and FIBTEM A5 levels observed in cohort

In total 637 valid combinations of measurements of Clauss fibrinogen and FIBTEM levels were obtained. Median FIBTEM A5 level was 17mm (IQR 13 to 20) and median Clauss fibrinogen concentration 3.9 g/L (IQR 3.1 to 4.5). Of the 511 women, 31 (6%) had a Clauss fibrinogen concentration of ≤2 g/L.

### Correlation Clauss fibrinogen ROTEM® FIBTEM A5

In the 637 samples, overall correlation between Clauss fibrinogen concentration and FIBTEM A5 showed a Spearman’s correlation coefficient (r_s_) of 0.64 (95% Confidence Interval (CI): 0.60 to 0.69). Spearman’s correlation coefficient between Claus fibrinogen concentration and FIBTEM measured on the ROTEM® Delta and Sigma device were (r_s_) 0.63 (95% CI: 0.58 to 0.67) and (r_s_) 0.76 (95% CI: 0.63 to 0.85) respectively (Figure 2). Within the three sample categories, Spearman’s correlation coefficients improved with increasing volumes of blood loss (Table 2).

**Figure 2:**
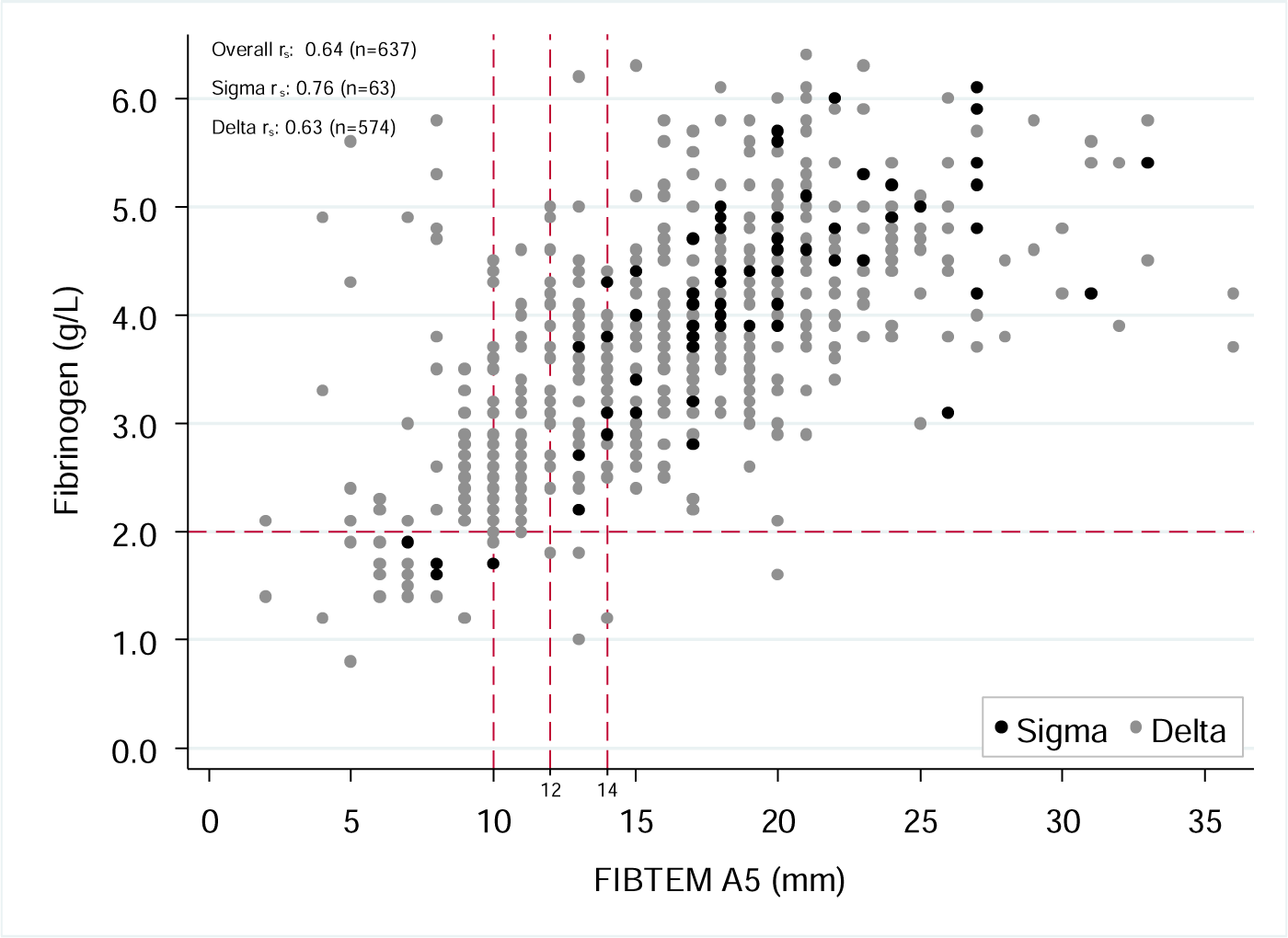
scatterplot correlation Clauss fibrinogen and FIBTEM A 5

**Table 2:** Fibrinogen concentration as assessed with the Clauss assay, ROTEM® FIBTEM A5 values and their correlations according to increasing volumes of blood loss among 511 women with postpartum haemorrhage.

| *Samples* | *Volume of blood loss\* (L)* | *Clauss Fibrinogen\* (g/L)* | *FIBTEM A5\* (mm)* | *Spearman's correlation (r<sub>s</sub>) (95% CI) †* |
| --- | --- | --- | --- | --- |
| **1.0-1.5 L (n = 412)** | 1.0 (0.9 to 1.2) | 4.1 (3.5 to 4.7) | 18 (14 to 20) | 0.56 (0.48 to 0.62) |
| **1.5-2.5 L (n = 185)** | 1.8 (1.5 to 2.0) | 3.5 (2.9 to 4.1) | 15 (12 to 18) | 0.67 (0.58 to 0.74) |
| **> 2.5 L (n = 40)** | 2.7 (2.5 to 3.4) | 2.7 (2.2 to 3.3) | 12 (10 to 16) | 0.79 (0.64 to 0.89) |
| **All samples (n = 637)** | 1.2 (1.0 to 1.6) | 3.9 (3.1 to 4.5) | 17 (13 to 20) | 0.64 (0.60 to 0.69) |

\* Values in the table reported in this format are median and (IQR),
† Spearman's correlation (r<sub>s</sub>) (95% CI)

### Discriminative ability of FIBTEM A5 for low fibrinogen concentration and cut-off points

The ability of FIBTEM A5 to select observations with a Clauss fibrinogen concentration of ≤2 g/L was good, area under Receiver Operating Curve 0.92 (95% confidence interval (CI) 0.87 to 0.97). Based on the Youden index, the best cut-off point in our cohort was 12mm with a sensitivity of 87% and specificity of 81%, missing 4 of the 31 cases (13%) of a Clauss fibrinogen ≤2 g/L and false positively selecting 118 cases (81%) (Table 3). When 15 mm was used as FIBTEM A5 cut-off point, 97% (30/31) of the samples with a Clauss fibrinogen ≤2 g/L were accurately selected, but 89% (248/278) of selected samples had a fibrinogen concentration >2 g/L. When a lower cut-off value for FIBTEM A5 was chosen, these numbers changed: a cut-off value of FIBTEM A5 of 6 mm accurately selected 26% (8/31) of samples with a Clauss fibrinogen concentration of ≤2 g/L and 74% (23/31) of samples with a low fibrinogen were missed. Yet, a lower percentage of 53% (9/17) of samples that were selected based on FIBTEM A5 value < 7 mm had corresponding Clauss fibrinogen values of >2g/L.

**Table 3.** Diagnostic test characteristics for FIBTEM A5 at various cut-off values to detect Clauss fibrinogen ≤2g/L.

| FIBTEM (mm) | Sensitivity | Specificity | Proportion of samples with fibrinogen $\leq 2$ g/L incorrectly classified as fibrinogen $> 2$ g/L* | Proportion of samples with FIBTEM A5 below cut-off, incorrectly classified fibrinogen $\leq 2$ g/L† |
| --- | --- | --- | --- | --- |
|  |  |  | False negative | False positive |
| A5 $\leq 6$ | 26% | 99% | 74% (23/31) | 53% (9/17) |
| A5 $\leq 7$ | 52% | 98% | 48% (15/31) | 43% (12/28) |
| A5 $\leq 8$ | 65% | 97% | 36% (11/31) | 50% (20/40) |
| A5 $\leq 9$ | 68% | 94% | 32% (10/31) | 63% (36/57) |
| A5 $\leq 10$ | 77% | 90% | 23% (7/31) | 71% (60/84) |
| A5 $\leq 11$ | 81% | 85% | 19% (6/31) | 79% (92/117) |
| A5 $\leq 12$ | 87% | 81% | 13% (4/31) | 81% (118/145) |
| A5 $\leq 13$ | 94% | 73% | 6% (2/31) | 85% (162/191) |
| A5 $\leq 14$ | 97% | 67% | 3% (1/31) | 87% (202/232) |
| A5 $\leq 15$ | 97% | 59% | 3% (1/31) | 89% (248/278) |

\* In the numerator the number of women with a FIBTEM at or below the cut-off and a fibrinogen concentration $\leq 2$ g/L, in the denominator the total number of women with a fibrinogen concentration $\leq 2$ g/L.
† In the numerator the number of women with a FIBTEM at or below the cut-off and a fibrinogen concentration $> 2$ g/L, in the denominator the total number of women with a FIBTEM at or below the cut-off.

## Discussion

Based on the Youden index, the optimal cut-off point in our cohort was a FIBTEM A5 of 12mm. If FIBTEM A5 would have been used to detect women with a low fibrinogen concentration in a treatment algorithm that uses the 12 mm cut-off to initiate administration of fibrinogen concentrate, 87 percent of women in need of fibrinogen would have been treated. However, a large majority of women (81%) would have received fibrinogen concentrate despite normal plasma fibrinogen concentrations.

## Strengths and limitation

To the best of our knowledge this is the largest study to date to describe fibrinogen concentrations according to previously proposed FIBTEM A5 cut-off points in blood samples collected from women suffering postpartum haemorrhage. The strength of this prospective study is that we were able to include a large cohort of women sustaining postpartum haemorrhage with repeated blood samples containing set panels at established times during postpartum haemorrhage. This enables for reliable and generalizable estimation of correlation and determination of interventional cut-off points based on Clauss fibrinogen and FIBTEM A5 levels. The prospective design of our study also has limitations: although the majority of women consented to participate in the study during their pregnancy, or in case they did not give consent yet, the opportunity was created to provide verbal consent during early postpartum haemorrhage, we did experience the challenge of performing study procedures in acutely ill women as described by others^15^. A total of 31 (6.1%) of the 511 women in our cohort developed a Clauss fibrinogen concentration ≤2g/L. Keeping track of missed cases that could have been included in the study, we noticed that women who lost a large volume of blood in a short period of time were more frequently not included in the study due to lack of time and personnel to perform study procedures in such an acute situation. This may have led to an underestimation of the incidence of a Clauss fibrinogen concentration ≤2g/L. However, the incidence of a Clauss fibrinogen concentration ≤2g/L in our source population of women giving birth during in the participating hospitals during the inclusion period was 0.2% (31/17203) which is exactly the same (0.2%, 342/191772) as observed in a previous study with the same source population (women giving birth in the Netherlands)^10^.

## Comparison with other studies

Correlation between Clauss fibrinogen concentration and FIBTEM A5 in women experiencing postpartum haemorrhage has been described in two previous studies. One observational study with postpartum haemorrhage (>500mL after vaginal birth or >1000mL after caesarean section) found a Spearman’s correlation coefficient (r_s_) of 0.86 for the haemorrhage group and r_s_ 0.83 for the control group^22^. No information was provided with regard to volume of blood loss at the time of sampling and number of samples with a Clauss fibrinogen ≤2g/L, yet correlation coefficients were similar for both groups, whereas we found an increase in correlation with higher volumes of blood loss. Another study into FIBTEM A5 as a biomarker for progression of postpartum haemorrhage found a moderate correlation in 312 paired Clauss fibrinogen and FIBTEM A5 assays of r 0.59^13^. These assays were sampled at study entry, at a median volume of blood loss of 1200mL. Our findings corroborate these results with a r_s_ of 0.55 at 1000mL and r_s_ of 0.67 at 1800mL. The choice for a cut-off point for a diagnostic test depends on the risks for adverse outcomes when patients are incorrectly classified and/or incorrectly treated. Since the risk of adverse outcomes is very high when fibrinogen concentrations are below 2 g/L, sensitivity of the test to diagnose all women with fibrinogen below 2 g/L should be high. At the time of writing, there is no consensus on an optimal cut-off point for FIBTEM A5 and results of previous studies lack conformity in their conclusions. In the previously cited study with high correlation between FIBTEM A5 and Clauss fibrinogen, a FIBTEM A5 level of 6 mm was found to correspond best (sensitivity 100%, specificity 87%) to the threshold of Clauss fibrinogen ≤2g/L^22^. Yet, in other studies, a FIBTEM A5 level of 12mm and 15 mm were considered the equivalent of a Clauss fibrinogen concentration of 2 and 3 g/L respectively^13, 15, 23^. No additional explanation was provided to support the choice for these thresholds. In a double-blind randomised controlled trial (OBS2), the effect of early fibrinogen replacement in women during postpartum haemorrhage guided by thromboelastometry was examined^15^. In this study women, with a FIBTEM A5 value of ≤15mm were randomised to treatment with fibrinogen concentrate or placebo based on an earlier observational study of the same research group, which found that a FIBTEM A5 value of ≤ 15mm was associated with progression of postpartum haemorrhage. No reduction of allogeneic blood product transfusion or volume of blood loss was observed. Also, a subgroup analysis in women with a FIBTEM A5 value of ≤ 12mm showed no significant differences between groups. However, in this study only seven women developed a Clauss fibrinogen concentration ≤2 g/L.

## Clinical implications

During acute situations like postpartum haemorrhage, clinicians are keen to get fast, reliable results on a woman’s coagulation status. FIBTEM A5 has been promoted to diagnose fibrinogen deficiency and guide treatment with fibrinogen concentrate. Assuming that women suffering postpartum haemorrhage with a Clauss fibrinogen concentration of ≤2 g/L require administration of fibrinogen concentrate, FIBTEM A5 is useful but lacks specificity. Using FIBTEM A5 with a cut-off point of 12 mm will lead to large numbers of women receiving fibrinogen concentrate in vain. The development of a point-of-care test that accurately and rapidly measures fibrinogen concentrations could be of considerable clinical significance.

## Conclusion

Based on the Youden index, the best cut-off point to accurately select women with a Clauss fibrinogen concentration of ≤2 g/L is FIBTEM A5 of 12mm. Yet, treatment of all women with FIBTEM A5 lower or equal than 12 mm with fibrinogen concentrate will lead to inappropriate use of fibrinogen in about 80 percent of women with postpartum haemorrhage treated with fibrinogen concentrate.

## Data Availability

All data produced in the present work are contained in the manuscript

## Acknowledgements

We would like to thank the women who participated in the TeMpOH-2 study, scientific laboratory technician D. Priem- Visser, research nurses C. Kolster-Bijdevaate, M.S. Bourgonje-Verhart, C.E. Bleeker-Taborh, E. Roos-van Milligen, R.J. M. Berkhout, E. Sucu, E. C. Willems of Brilman-Tuinhof de Moed and M. Stigter-Dekker and medical students M. van de Sande, R.H. Wouters, and L.S. Smits and the clinical midwifes of the Leiden University Medical Center for their contributions to the TeMpOH-2 study.

## Contribution to authorship

Contribution: A.G., J.B. designed the research and A.G. wrote the original draft of the paper. A.G and C.C. were responsible for data curation. C.C., B.G. and A.G. analysed results and made the figures and tables. A.G.,D.H., H.D., M.M., J.E.,K.B.,J.B. were involved in conceptualization and methodology. All co-authors reviewed and edited the paper and gave final approval. J.B. and T.A. had supervision over the project.

## Details of Ethics Approval

Approval for the study was obtained from the Ethical Committee of the Leiden University Medical Centre (P13.246) and from the institutional review board of each participating hospital. The study was registered at ClinicalTrials.gov (NCT02149472).

